# *VAREANT*: a bioinformatics application for gene variant reduction and annotation

**DOI:** 10.1101/2024.09.24.24314291

**Authors:** Rishabh Narayanan, William DeGroat, Elizabeth Peker, Zeeshan Ahmed

## Abstract

The analysis of high-quality genomic variant data may offer a more complete understanding of the human genome, enabling researchers to identify novel biomarkers, stratify patients based on disease risk factors, and decipher underlying biological pathways. Although the availability of genomic data has sharply increased in recent years, the accessibility of bioinformatic tools to aid in its preparation is still lacking. Limitations with processing genomic data primarily include its large volume, associated computational and storage costs, and difficulty in identifying targeted and relevant information. Here, we present *VAREANT*, an accessible and configurable bioinformatic tool to support the preparation of variant data into a usable analysis-ready format. Designed to simplify the data pre-processing workflow, *VAREANT* enables the curation of targeted variant datasets. *VAREANT* is comprised of three standalone modules: (1) Pre- processing, (2) Variant Annotation, (3) Artificial Intelligence (AI) / Machine Learning (ML) Data Preparation. Pre-processing supports the fine-grained filtering of complex variant datasets to eliminate extraneous data. Variant Annotation allows for the addition of variant metadata from public annotation databases for subsequent analysis and interpretation. AI/ML Data Preparation supports the user in creating AI/ML-ready datasets suitable for immediate analysis with minimal pre-processing required. We have successfully tested and validated our tool on numerous variable-sized datasets and implemented *VAREANT* in two case studies involving patients with CVDs. Efficiently extracting relevant variants into an AI/ML ready format using tools like *VAREANT* has important scientific implications, such as producing targeted and high-quality datasets and helping reduce overall computational costs.

## Introduction

Advancements in genome sequencing technologies have resulted in an immense wealth of available genomic data. The analysis of genetic variations via genome-wide association studies (GWAS) can improve our understanding of disease prognosis, treatments, and etiology by helping to uncover disease-causing variants and complex gene-disease relationships [1]. Recent progress in artificial intelligence (AI) and machine learning (ML) techniques have demonstrated their efficacy for genomic predictive analysis [2]. The introduction of AI/ML prediction tools in the field of genomics has increased our understanding of disease etiology and shows potential for further uptake in clinical practice [3]. However, the rapid growth of genomic data presents many analytic obstacles. Most genomic data formats are not immediately suitable for AI/ML analyses, requiring extensive preprocessing. Although state-of-the-art bioinformatic tools have been created to individually support the various data transformation stages, there is a lack of tools to extract highly targeted genomic data suitable for immediate analysis. In addition, large volumes of heterogenous genetic information make data preparation difficult for targeted studies investigating only specific genes of interest. These datasets often include thousands of irrelevant data points resulting in wasteful processing and lengthy computation times [4]. Moreover, such workflows are often inaccessible to clinicians and translational researchers that lack the computational expertise [5, 6]. Ultimately, addressing these challenges will help render genomic analyses more accessible, affordable, and effective.

Processing variant data is an arduous process with many stages: raw sequence files undergo many transformations such as quality checking, trimming, alignment to a reference genome, variant calling, filtering, annotation, analysis, and visualization. Many tools exist to streamline each of these stages, including Burrows-Wheeler Aligner (BWA) [7] for alignment, Genome Analysis Toolkit (GATK) [8] for variant calling, and SnpEff [9] or Ensembl Variant Effect Predictor (VEP) [10] for annotation. To support the variant calling workflow, we have recently developed a reliable Java-based Whole Genome/Exome (JWES) Data Processing Pipeline [11]. JWES centralizes these various tools into a single cohesive pipeline for processing variant files. Using BWA for alignment, GATK for variant calling, and SnpEff for annotation, JWES allows the user to easily prepare a Variant Call Format (VCF) dataset. The VCF format is an extensible data format to encode mutation data alongside corresponding annotations [12], and it is widely supported by many genomic tools. To help address these data processing challenges, we have developed **<u>VA</u>riant <u>RE</u>duction and <u>AN</u>noTation** (*VAREANT*), a configurable and accessible bioinformatic tool to support the curation of targeted variant and AI/ML-ready datasets. *VAREANT* is designed as a series of standalone modules to support the user with their various data preparation needs. It was validated in a case study with a selective cohort of patients with cardiovascular diseases (CVDs) to extract relevant CVD-associated variants and annotations. With its utilization in data management pipelines, *VAREANT* can help simplify the data transformation workflow and produce effective datasets equipped for subsequent AI/ML analyses.

## Materials and methods

*VAREANT* is split into three standalone modules: I) Preprocessing, II) Variant Annotation, and III) AI/ML Data Preparation. Each module may be used independently on custom datasets or chained together to robustly extract relevant variant data from a larger genomics dataset.

### Preprocessing

To reduce overall costs associated with genomic data management and analysis, it is important to successfully identify and minimize any extraneous data. To support the curation of these targeted genomic datasets, *VAREANT* implements a highly efficient and customizable filtering methodology for selecting maximally relevant variants and metadata. Through the application of different filters, the user has nuanced control over which data points are retained, including genes, quality scores, pathogenicity scores, sample genotype data, annotations, allele frequencies, etc. Notably, *VAREANT* enables the selective extraction of both variants using rsIDs and genes by either gene symbols or Ensembl IDs [13]. This is particularly useful when the target gene set is known a priori, such as in many clinical studies. In addition to extracting gene-related data, specific variant metadata important for subsequent analysis may also be retrieved. Heavily annotated variant files may contain hundreds of features per variant, amassing to large volumes of potentially unused data. Filtering these extraneous annotations can lead to drastic reduction of file sizes. Variant files may also contain information about multiple sequenced samples, such as read depth, read quality, or haplotype phasing. For targeted analyses or case/control studies where only a subset of this data is required for investigation, *VAREANT* can efficiently extract this information. *VAREANT* was developed to be performant in different environments with variable resources, by efficiently taking full advantage of the available computing hardware. Larger files are split into smaller manageable chunks and processed efficiently on multiple central processing units (CPUs) in parallel. Moreover, by dynamically streaming chunks into memory, *VAREANT* can maintain a minimal memory footprint. We have open sourced our command line tool, written in Python, and made it publicly available on our GitHub. With its simple interface, it is accessible to researchers and clinicians lacking a computational background, requiring only a basic understanding of executing scripts. Details on installing and using *VAREANT* can be found in the Supplementary User Guide.

### Variant Annotation

Variant annotation is the process of associating metadata from public databases with corresponding variant data. The choice of annotations can have a significant impact on the interpretation and conclusions of genomic analyses [14]. Although many other annotation tools exist, such as VEP or ANNOVAR, *VAREANT* employs SnpEff due to its simplicity, portability, and efficiency [9]. For annotation databases, *VAREANT* currently supports dbSNP, dbNSFP, and ClinVar to identify clinical significance and functional pathogenicity scores for known variants. dbSNP is a public and centralized repository of genetic variation constituting primarily of single nucleotide polymorphisms (SNPs), the most common type of genetic variation [15]. However, dbSNP makes no distinction between neutral and pathogenic variants. For this purpose, dbNSFP can be used to provide 36 different functional pathogenicity scores aiding in determining variant deleteriousness [16]. For example, SIFT and PolyPhen scores may be used to predict whether a mutation is likely to affect protein structure, or Combined Annotation-Dependent Depletion (CADD) [17] and Eigen scores [18] which integrate numerous functional annotations to generate a single ML-based deleterious metric. Effective pathogenicity scores are crucial for identifying variations of interest and discovering disease etiology. Lastly, ClinVar is a public archive of genetic variants and their significance in human disease [19] and can be used as a basis for investigating variant-disease associations. Using *VAREANT*, the user may optionally annotate their datasets with any of these public databases depending on the study-specific needs. Since variant annotation is more time consuming for larger files, the user may pre-process their dataset using *VAREANT* before annotation to reduce the overall processing burden. Moreover, to support and enable scientists with their own custom annotation pipelines, the user can integrate *VAREANT’s* standalone modules independently with their own tools and annotation databases, widening the scope of use cases for *VAREANT*.

### AI/ML Data Preparation

AI/ML techniques show promise in progressing modern genomic analysis and personalized treatment by aiding scientists in understanding the genetic basis of disease [2, 20]. However, extensive preprocessing is often required to prepare data in a format conducive to AI/ML analysis. Although the VCF format is effective at encoding variation data, it is not suitable for immediate use. To prepare variant files for subsequent AI/ML analysis, *VAREANT* supports the extraction and transformation of variant and sample data into a tabular AI/ML-ready structure. This tabular structure is preferred over other formats for AI/ML analysis. It enables the integration of clinical demographic information with genomic data and is well supported by various programming languages. In addition to a tabular format, we recognize that bioinformaticians often have their own data management pipeline. To better support these custom workflows, the user may also extract data into a JWES-compatible relational database [11], enabling them to integrate their datasets with existing structured query language (SQL) data management solutions. This relational structure is more convenient for organizing variant annotations with sample data. *VAREANT* was written in the Python programming language and requires at least Python 3.6 to be installed on the system. It is compatible with all major operating systems including Windows, MacOS, and Linux. For annotation, *VAREANT* uses SnpEff which requires Java to be installed on system. For AI/ML data preparation, the user must also install the Pandas and NumPy python packages. We have open-sourced our tool to be accessible to the wider scientific community. Further installation and usage instructions can be found on our public GitHub as well as in our Supplementary User Guide.

## Results

### Data Collection

*VAREANT* has been successfully tested and validated in-house on varied datasets and environments. We carefully crafted 97 datasets of varying sizes: 96 samples were sequenced from separate patients with different CVDs, and the final dataset was curated from the 96 samples to simulate a more variant-dense dataset. The 96 samples were aggregated from two peer-reviewed studies investigating impactful genes and their associations with CVD, including atrial fibrillation (AF) and heart failure (HF). The first cohort contained 61 patients [21], and the second contained the remaining 35 [22]. Combined, 61 patients were male and 35 were female, aged between 24 and 94. Age and gender information for each patient is enumerated in the Supplementary Materials. *VAREANT* was used to preprocess and annotate each of these datasets ranging from 0.5 gigabytes (GB) to 3.1 GB in size. To further validate *VAREANT*, we derived a literature-based set of CVD-associated genes and variants to cross-reference the extracted results. We recently conducted a thorough review of literature published between 2009 and 2022 focused on integrative genomic approaches, common and rare genetic variant analyses for CVDs, and multi-ethnic studies [23]. In that study, we identified a total of 214 variants from 190 genes associated with AF, and 28 variants from 26 genes associated with HF. This gene set of 216 genes was used as the primary criterion for filtering and annotation with *VAREANT*. All filtered variants were manually reviewed to ensure consistency with the specified gene set. A list of variants from our criterion that were identified in our dataset may be found in the Supplementary Materials. The original 96 samples consisted of 396,788,923 total variants, which was narrowed down by over a factor of 100 to 3,906,744 variants using *VAREANT*. Annotation on this targeted dataset was subsequently performed using dbSNP, dbNSFP, and ClinVar. dbSNP (dated 04/23/2018) was used to identify each variant by their rsIDs, and dbNSFP (v4.1a) and ClinVar (dated 07/08/2024) was used to determine pathogenicity and clinical significance of each variant.

### Case Study #1: Heart Failure

From our processed dataset, we first identified the total number of variants on genes associated with HF. Specifically, 220,348 variants of the 3.9 million variants filtered by *VAREANT* belonged to HF-associated genes, according to our filtering criterion. We then explored the individual annotations to identify variants with known clinical significance and pathogenic effects. 12,373 of these variants reported clinical significance and pathogenicity annotations, most of which were marked benign or with uncertain significance. The remaining novel variants lacked any annotations. In total, 9 unique variants were successfully identified from ClinVar annotations as having some pathogenic association in a GWAS study or being a risk factor for disease. Specifically, deleterious variants were labelled as either ‘pathogenic,’ ‘likely pathogenic,’ ‘risk factor,’ or ‘association,’ as described by ClinVar’s variant classification guidelines [19]. Specifically, *rs1063192* was indicated to be likely pathogenic; *rs977371848, rs12740374, rs947073006* were marked as having some association to GWAS a study; and *rs1333049, rs10757274, rs1421085, rs4977574* were identified as being risk factors. From our original set of 28 HF-associated variants, 21 were present in our dataset and all were marked benign/likely benign. These variants are enumerated in the Supplementary Materials.

To determine the significance of each variant in HF and CVDs in general, we reviewed authentic literature. We discovered that *rs1063192* has been studied to have a positive association with myocardial infarction (MI) in Han Chinese male patients [7]; *rs1421085* has associations with childhood and adult obesity [24, 25]; *rs1333049* has been studied to have associations with coronary artery disease (CAD) in Caucasian [26], Japanese [27], and Korean populations [27], and associations with MI in German populations [28]; *rs4977574* shows change in African and Middle-Eastern populations for type 2 diabetes and CAD [29]; *rs10757274* has been previously associated with MI in Italian [30], as well as CAD in Korean populations

[31]; *rs12740374* was studied to be highly associated with low-density lipoprotein cholesterol [32]. The remaining variants were not previously studied to have associations with HF. An aggregate of these variants, their prevalence in our dataset, and their clinical significance is enumerated in Table 2.

**Figure 1.**
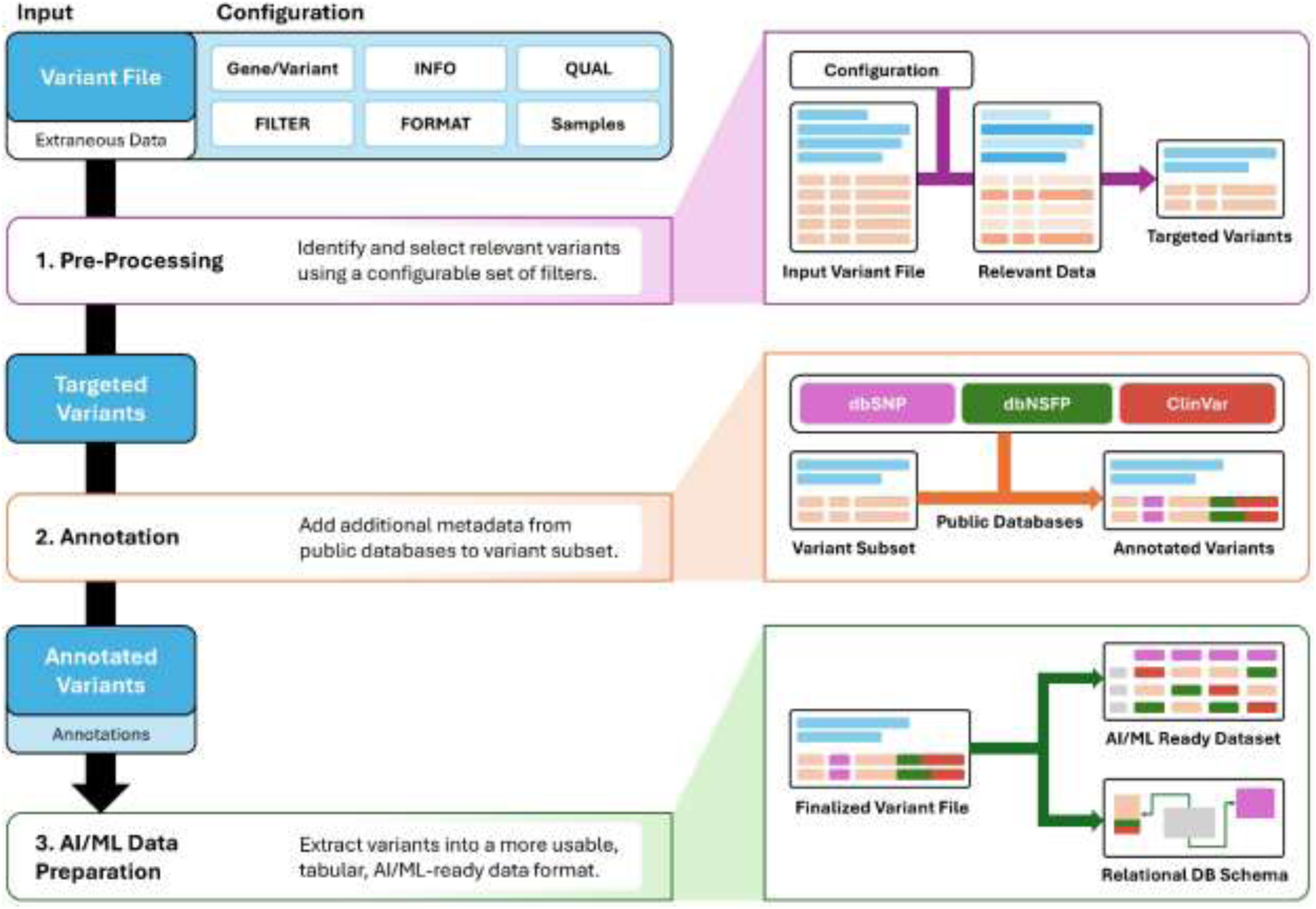
*VAREANT* pipeline overview (left) and data transformation workflow (right): 1) Pre-Processing supports user in curating targeted datasets, 2) Annotation supports user in easily annotating variants files, 3) AI/ML Data Preparation supports user in extracting VCF files into a more usable AI/ML ready data format.

**Table 1.**
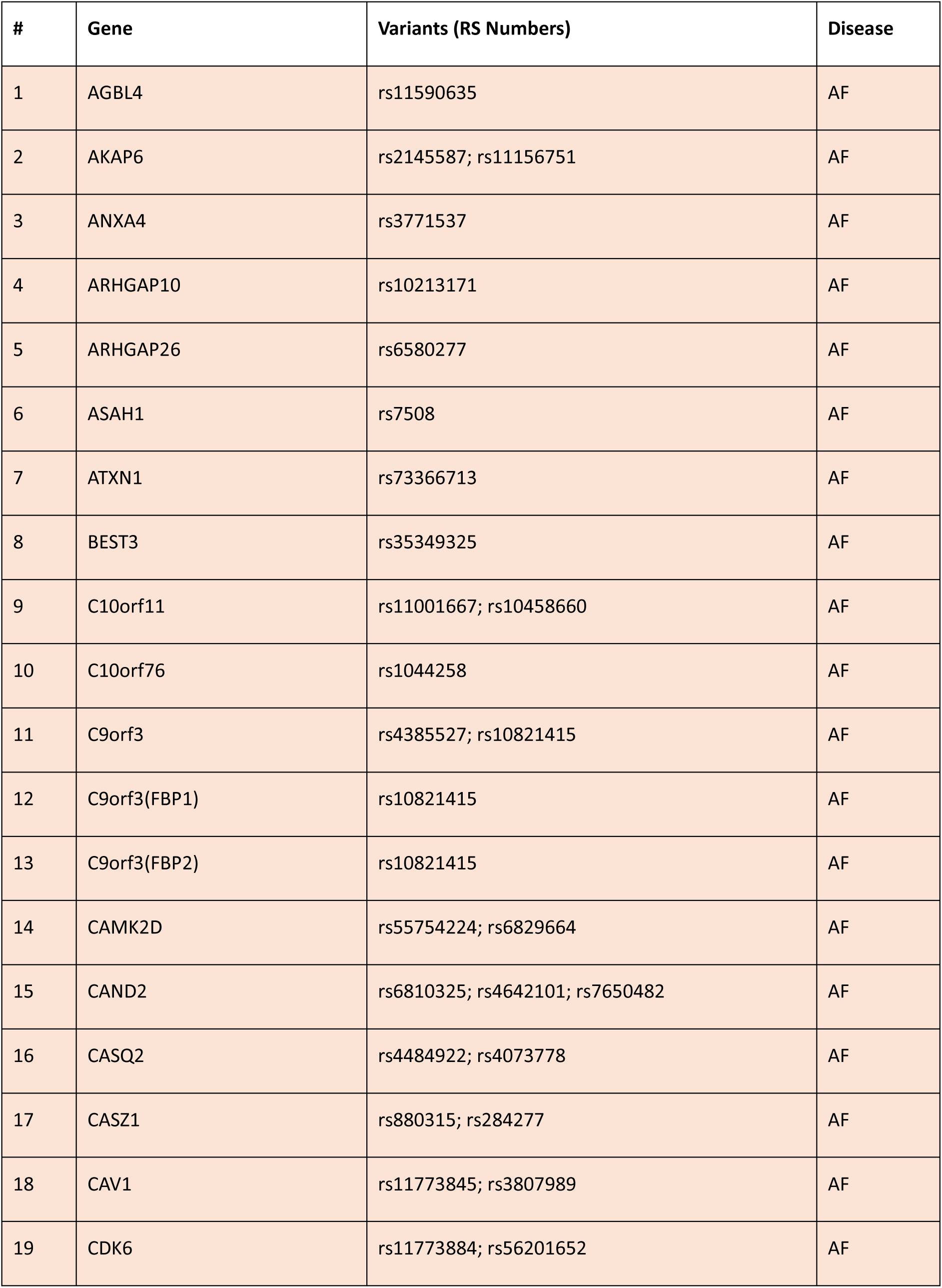
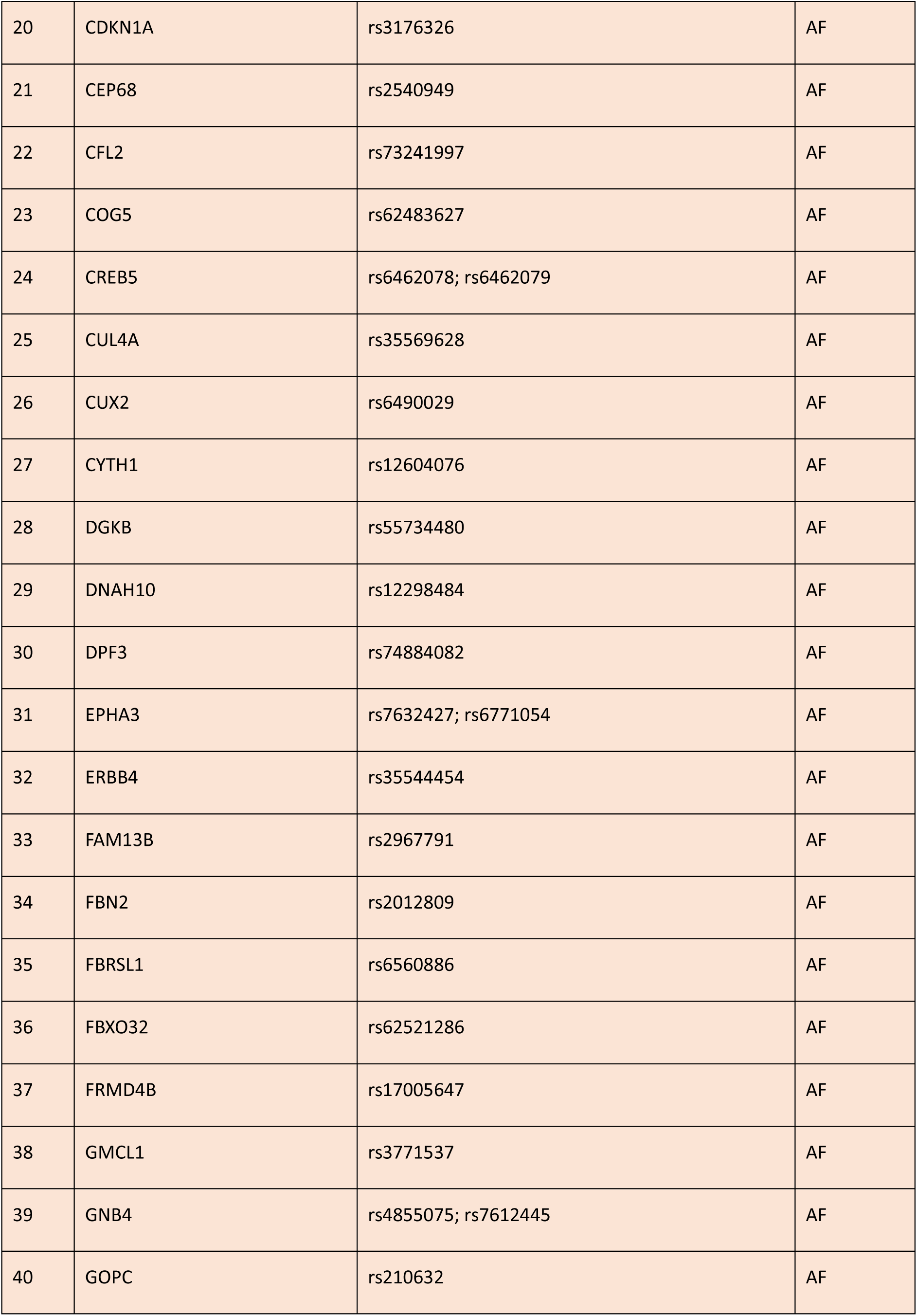
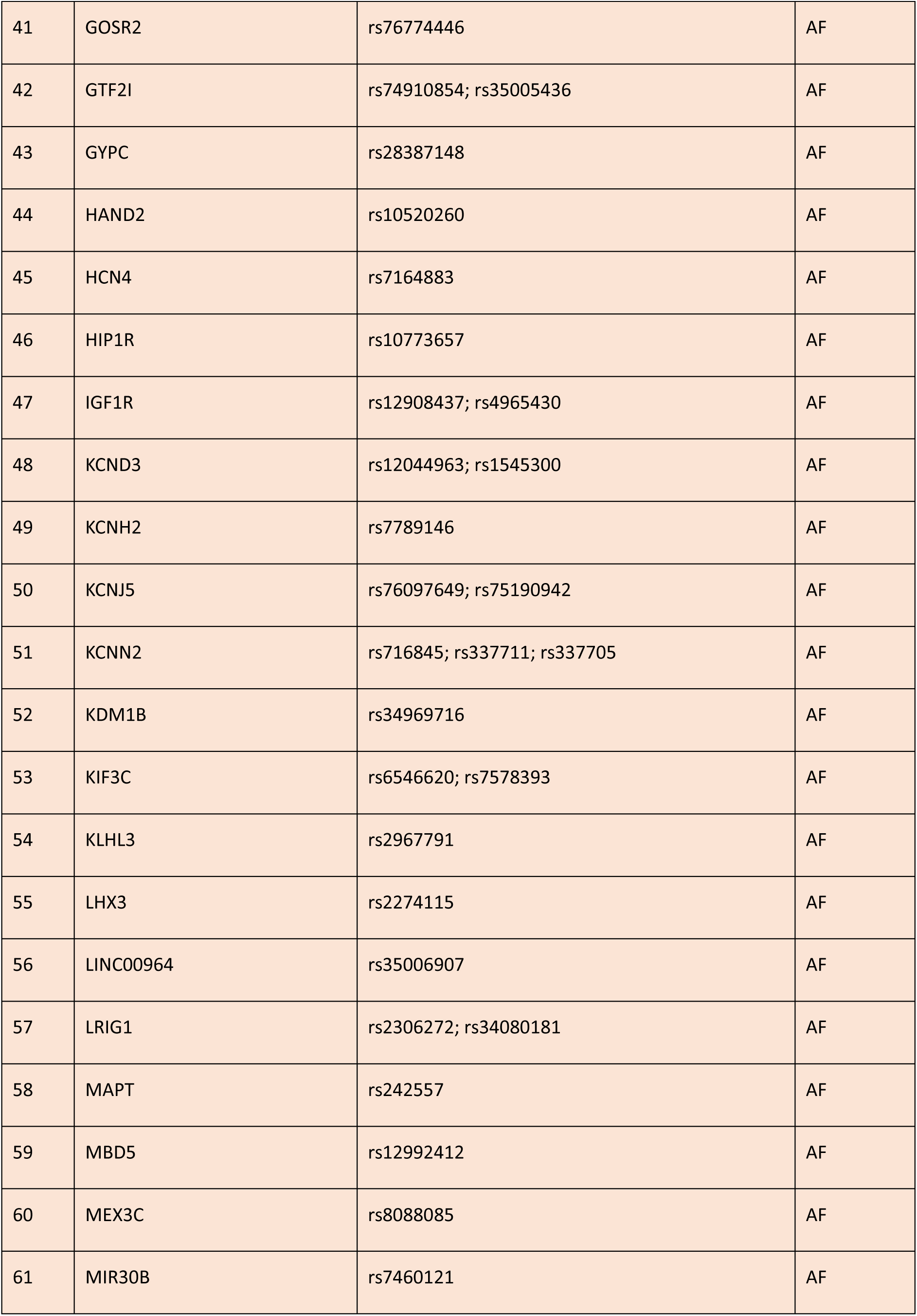
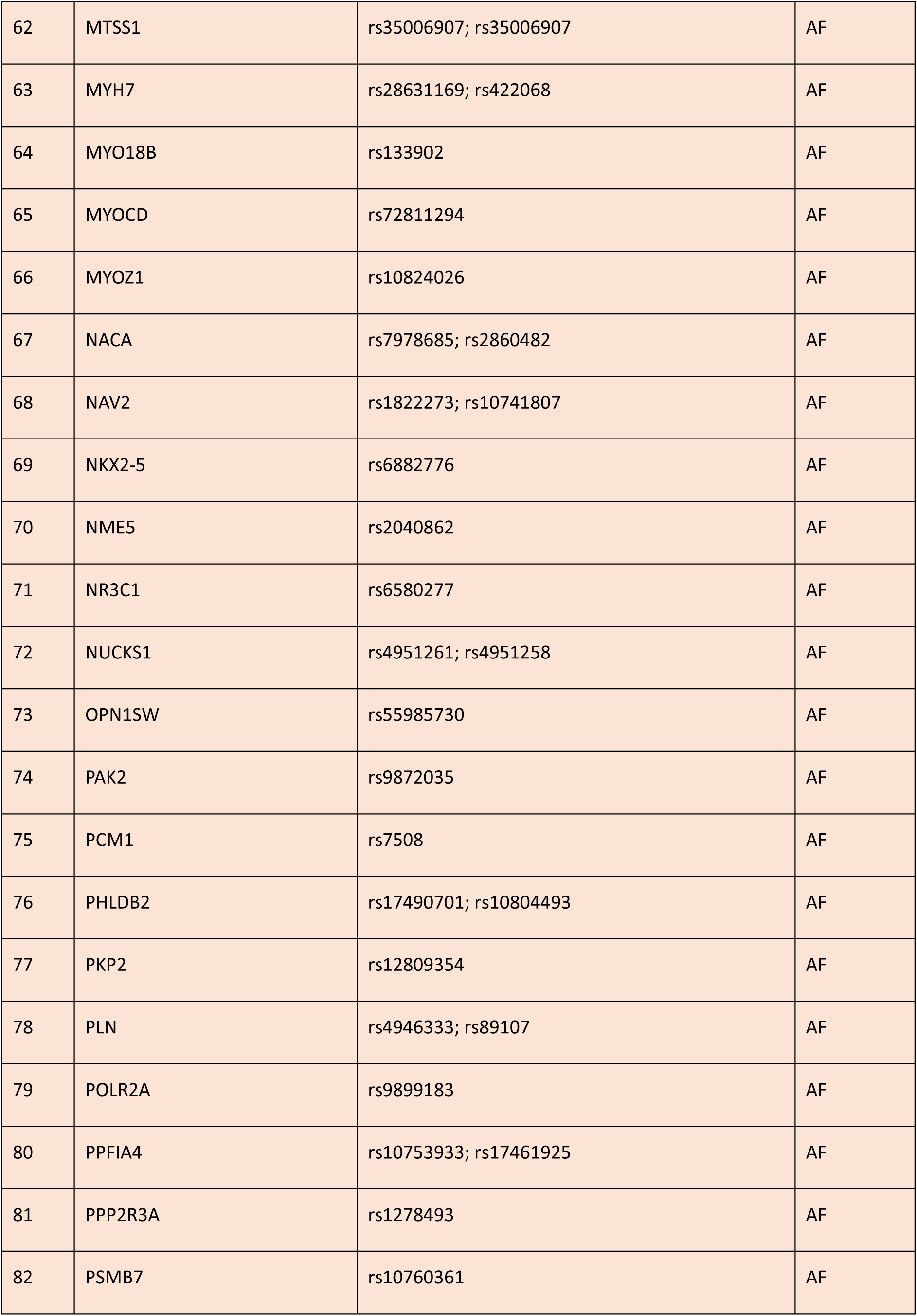
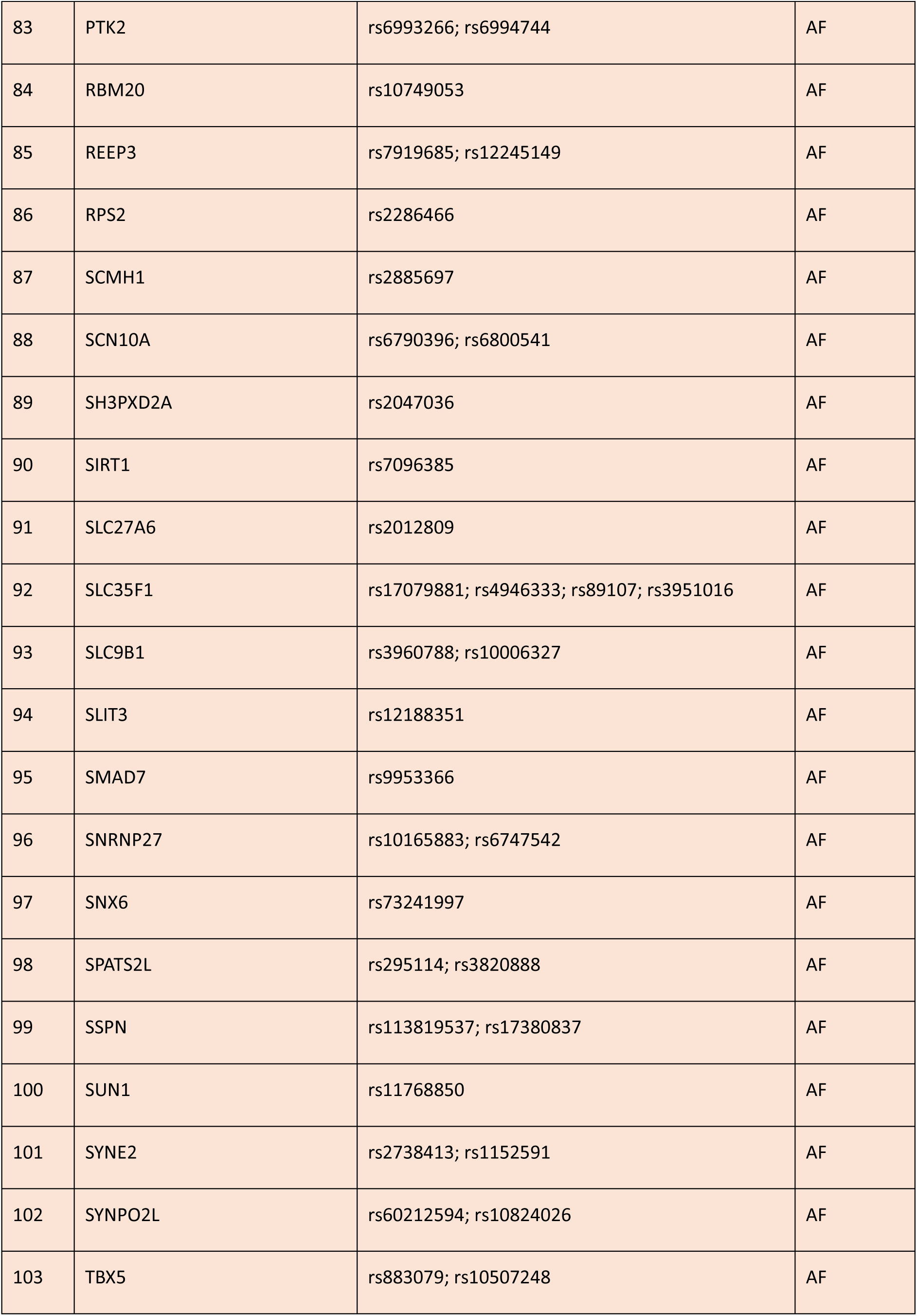
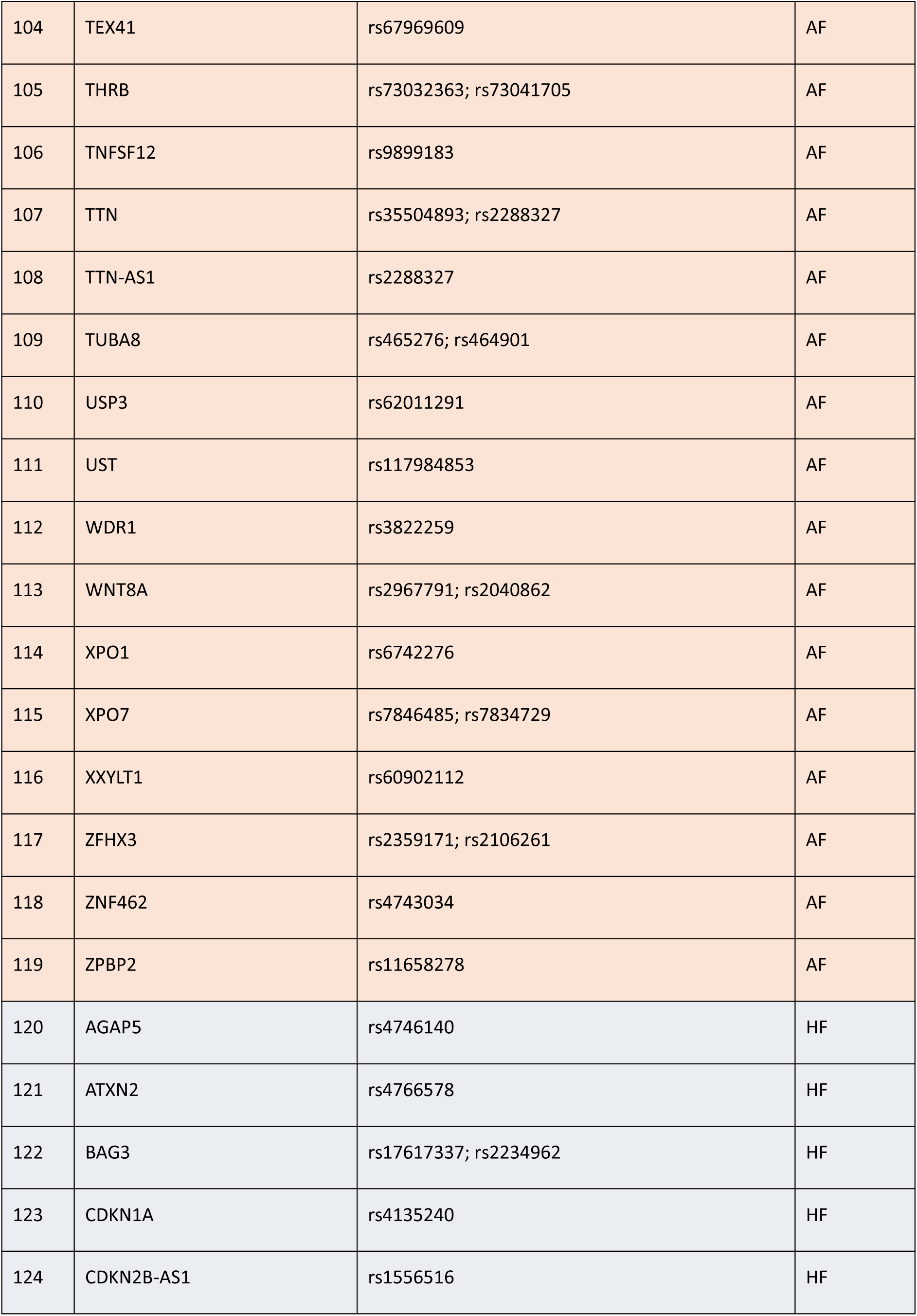
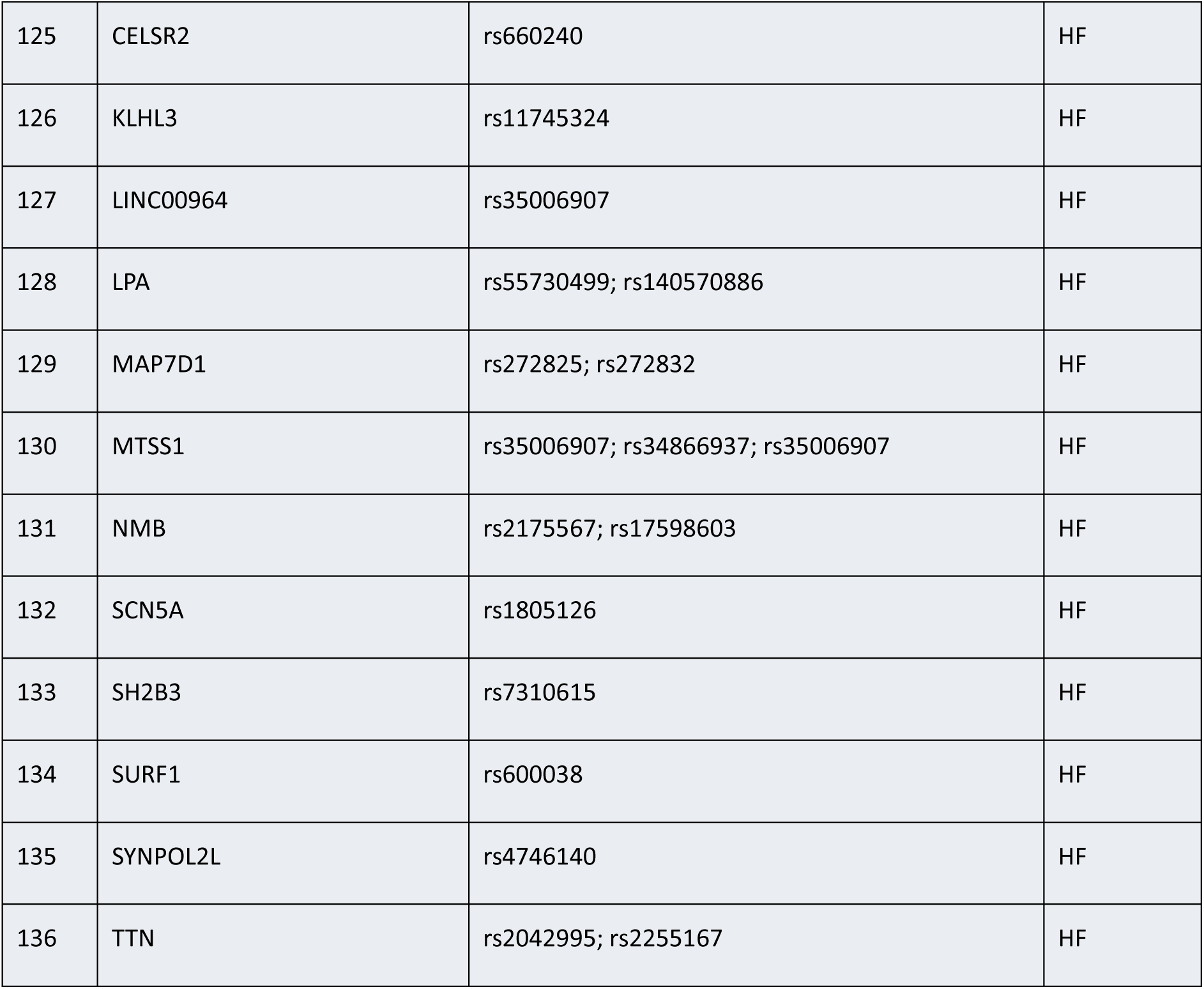
Gene-Variants Associated with AF/HF. This table includes genes, variant (RS Numbers), and Disease information. Genes associated with Atrial Fibrillation (AF) are colored orange, and those associated with Heart Failure (HF) are colored blue.

**Table 2.** Disease-Associated Variants Identified by ClinVar. This table enumerates genes, RS Number, frequency in cohort, pathogenic scoring based on ClinVar, and associated diseases for 16 variants. These variants were identified by ClinVar as having some known association to diseases.

| Gene | Variant (RS Number) | Frequency (in 96 sample cohort) | Clinical Significance based on ClinVar | Disease Names based on ClinVar |
| --- | --- | --- | --- | --- |
| COG5 | rs1449966934 | 1 | Pathogenic | COG5 Congenital Disorder of Glycosylation |
| CDKN2B-AS1 | rs1063192 | 76 | Likely Pathogenic | Malignant tumor of breast, Three Vessel Coronary Disease |
|  | rs1333049 | 70 | Risk Factor | Three Vessel Coronary Disease |
|  | rs4977574 | 70 | Risk Factor | Three Vessel Coronary Disease |
|  | rs10757274 | 71 | Risk Factor | Three Vessel Coronary Disease |
| FTO | rs1421085 | 70 | Risk Factor | Obesity (BMIQ14) Susceptibility |
| ABO | rs947073006 | 90 | Association | ABO Blood Group System |
|  | rs977371848 | 90 | Association | ABO Blood Group System |
|  | rs992108547 | 90 | Association | ABO Blood Group System |
| ARNT2 | rs3901896 | 55 | Association | Pulmonary Disease Susceptibility |
|  | rs8041826 | 31 | Association | Pulmonary Disease Susceptibility |
| CDK6 | rs42034 | 40 | Association | Bechet Disease |
|  | rs2282983 | 65 | Association | Bechet Disease |
| CELSR2 | rs12740374 | 32 | Association | LDL Cholesterol |
| CREB5 | rs4722804 | 30 | Association | Vascular Endothelial Growth<br>Factor Inhibitor Response |
| SLC35F1 | rs11153718 | 34 | Association | Vascular Endothelial Growth<br>Factor Inhibitor Response |

### Case Study #2: Atrial Fibrillation

Of the over 3.9 million variants filtered using *VAREANT*, the remaining 3,686,396 belonged to genes associated with AF. 101,745 of these variants were annotated to have some clinical significance, and 7 unique variants indicating some pathogenic clinical significance [19]. Namely, *rs1449966934* was marked pathogenic, and *rs8041826, rs4722804, rs2282983, rs11153718, rs3901896, rs42034* were all identified as having some previously studied GWAS association. From our original set of 214 AF-associated variants, 151 variants were present in our dataset, all of which were also annotated as benign/likely benign. These variants are enumerated in Table 2. To explore the relationship of the pathogenic variants with CVDs, we reviewed authentic literature for each variant. Notably, *rs42034* was studied to have negative associations with Bechet’s disease (an inflammatory disease) in the Han Chinese population [33]. The other 6 variants were not previously explored in literature, but *rs1449966934* was annotated to have associations with the congenital disorder glycosylation. Further study is required to identify any potential functional associations these variants may have with CVDs.

### Computational Validation

Performance was extensively tested and benchmarked on different operating systems (MacOS, Linux, Windows), on different hardware configurations (4 CPUs and 8 GB memory, 12 CPUs and 32 GB memory), in high performance computing (HPC) and local desktop environments, and in single-processor and multi- processor modes. The results of each configuration for the combined variant-dense dataset and for one of the 96 patient-specific datasets are listed in the Supplementary Materials. Notably, *VAREANT* successfully filtered out over 97% of the combined dataset (0.5 GB) and over 99.9% of the larger 2.6 GB patient-specific dataset within seconds. *VAREANT* also performs better on more powerful hardware, completing filtering nearly twice as fast on the larger dataset when using 12 CPUs over 4 CPUs. Moreover, on a single processor, limited memory does not appear to pose a noticeable bottleneck. The performance metrics also effectively delineate the impact that filtering with *VAREANT* has on subsequent annotation. Annotating the large 2.6 GB dataset using dbSNP, dbNSFP, and ClinVar took over 3 hours on an M2 MacBook, which was reduced to merely 16 seconds after first filtering the dataset for relevant variants only. The smaller yet variant-dense dataset also experienced significant improvements in annotation from nearly 15 minutes before filtering to only 44 seconds after filtering. Although these results are subject to the hardware resources and content of the dataset, we recommend running *VAREANT* in its multi-processor mode to take full advantage of provided hardware, and to filter before annotation to minimize computational times. This configuration is the default.

## Discussion

The curation of high-quality, relevant genomic datasets is necessary to facilitate genomic analysis that can transform our current understanding of gene-disease relationships and improve our ability to provide personalized treatment options for patients. Moreover, it may help minimize difficulties associated with genomic data management, such as the high storage capacities needed to handle large volumes of data [5] and the time-consuming processing [6]. In this study, we aim to address some of these challenges by presenting *VAREANT*, a highly configurable and accessible bioinformatics tool to process large volumes of variant data into targeted datasets suitable for subsequent analysis. Here, we demonstrated the efficacy of *VAREANT* in a case study of 97 CVD variant files. Using a literature-based gene set, we successfully created a targeted dataset and identified numerous variants with associations with CVD diseases, such as CAD and MI.

*VAREANT* was developed alongside a recent study we conducted wherein we applied AI/ML techniques to predict CVD in a patient population based on integrated RNA-Seq expression data and genomics variant data [34]. In that study, we faced significant obstacles in the processing and preparation of hundreds of GBs of genomics data in a format suitable for AI/ML analysis, prompting the need to curate a targeted, AI/ML-ready [35], CVD dataset. Although *VAREANT* was originally developed to support this specific study, we recognized the broader potential impact of such bioinformatic applications and developed it into a publicly available tool for the wider scientific community. Recently, we also developed and proposed *IntelliGenes*, a novel AI/ML framework for disease prediction and biomarker discovery in patients using multi-omics data [36, 37]. *VAREANT* is well suited to support the user with complex, multi-modal AI/ML analysis using tools like *IntelliGenes*. By chaining JWES, *VAREANT*, and *IntelliGenes*, the user can streamline the full genomic data transformation workflow. *VAREANT* may also be expanded to include more fine-grained filtering options for better data extraction. Here, we demonstrate that accessible bioinformatic tools such as *VAREANT*, to aid in preparing large volumes of data, are critical and effective in predictive genomic analyses.

## List of Abbreviations

AI: Artificial Intelligence
AF: Atrial Fibrillation
BWA: Burrows-Wheeler Aligner
CVD: Cardiovascular Disease
CPU: Central Processing Unit
CADD: Combined Annotation-Dependent Depletion
CAD: Coronary Artery Disease
VEP: Ensembl Variant Effect Predictor
GATK: Genome Analysis Toolkit
GWAS: Genome-Wide Assocation Studies
GB: Gigabytes
GUI: Graphical User Interface
HF: Heart Failure
HPC: High Performance Computing
JWES: Java-based Whole Genome/Exome Data Processing Pipeline
ML: Machine Learning
MI: Myocardial Infarction
SNP: Single Nucleotide Polymorphism
SQL: Structured Query Language
VCF: Variant Call Format
VAREANT: VAriant REduction and ANnoTation

## Acknowledgments

We appreciate great support by the Department of Medicine, Robert Wood John-son Medical School; Rutgers Institute for Health, Health Care Policy, and Aging Research; and Rutgers Health, at Rutgers, The State University of New Jersey.

## Author contributions

ZA proposed and led this study. RN developed VAREANT. WD supported its design and implementation. EP supported project management activities and post-computational analysis. RN also prepared the supplementary material. RN and WD tested and validated VAREANT. RN drafted the manuscript. All authors have reviewed and approved it for publication.

## Biographical Note

RN and EP are the Research Assistants, and WD is the Senior Research Assistant at the Ahmed lab, Rutgers IFH/RWJMS.

ZA is the Assistant Professor at the Department of Medicine / Division of Cardiovascular Diseases and Hypertension, Rutgers Robert Wood Johnson Medical School, and Rutgers Health. ZA is a Core Faculty Member at the Rutgers Institute for Health, Health Care Policy and Aging Research, at Rutgers, The State Universi-ty of New Jersey. Furthermore, ZA is the Adjunct Assistant Professor at the Department of Genetics and Genome Sciences, School of Medicine, UConn Health, CT.

## Data availability

The source code of *VAREANT* is available on GitHub < https://github.com/drzeeshanahmed/Gene_VAREANT >

## Supplementary materials

Supplementary Material 1: *VAREANT* User Guide.

Supplementary Material 2: Patient Demographic Data and Performance Metrics

## References

1. Uffelmann, E., Huang, Q. Q., Munung, N. S., de Vries, J., Okada, Y., Martin, A. R., Martin, H. C., Lappalainen, T., & Posthuma, D. (2021). Genome-wide association studies. Nature News. 10.1038/s43586-021-00056-9

2. Ahmed, Z., Mohamed, K., Zeeshan, S., & Dong, X. (2020). Artificial intelligence with multi-functional machine learning platform development for better healthcare and precision medicine. Database: the journal of biological databases and curation, 2020, baaa010. 10.1093/database/baaa010

3 Lin, Q., Tam, P. K., & Tang, C. S. (2023). Artificial intelligence-based approaches for the detection and prioritization of genomic mutations in congenital surgical diseases. Frontiers in pediatrics, 11, 1203289. 10.3389/fped.2023.1203289

4. Schadt E., et al. (2010). Computational solutions to large-scale data management and analysis. Nat Rev Genet 11, 647–657 (2010). 10.1038/nrg2857

5. Stephens, Z. D., Lee, S. Y., Faghri, F., Campbell, R. H., Zhai, C., Efron, M. J., Iyer, R., Schatz, M. C., Sinha, S., & Robinson, G. E. (2015). Big Data: Astronomical or Genomical?. PLoS biology, 13(7), e1002195. 10.1371/journal.pbio.1002195

6. Alvarez, R. V., Mariño-Ramírez, L., & Landsman, D. (2021). Transcriptome annotation in the cloud: complexity, best practices, and cost. GigaScience, 10(2), giaa163. 10.1093/gigascience/giaa163

7. Yang, X. C., Zhang, Q., Chen, M. L., Li, Q., Yang, Z. S., Li, L., Cao, F. F., Chen, X. D., Liu, W. J., Jin, L., & Wang, X. F. (2009). MTAP and CDKN2B genes are associated with myocardial infarction in Chinese Hans. Clinical biochemistry, 42(10-11), 1071–1075. 10.1016/j.clinbiochem.2009.02.021

8. McKenna, A., Hanna, M., Banks, E., Sivachenko, A., Cibulskis, K., Kernytsky, A., Garimella, K., Altshuler, D., Gabriel, S., Daly, M., & DePristo, M. A. (2010). The Genome Analysis Toolkit: a MapReduce framework for analyzing next-generation DNA sequencing data. Genome research, 20(9), 1297–1303. 10.1101/gr.107524.110

9. Cingolani, P., Platts, A., Wang, leL., Coon, M., Nguyen, T., Wang, L., Land, S. J., Lu, X., & Ruden, D. M. (2012). A program for annotating and predicting the effects of single nucleotide polymorphisms, SnpEff: SNPs in the genome of Drosophila melanogaster strain w1118; iso-2; iso-3. Fly, 6(2), 80–92. 10.4161/fly.19695

10. McLaren, W., Gil, L., Hunt, S. E., Riat, H. S., Ritchie, G. R., Thormann, A., Flicek, P., & Cunningham, F. (2016). The Ensembl Variant Effect Predictor. Genome biology, 17(1), 122. 10.1186/s13059-016-0974-4

11. Ahmed, Z., Renart, E. G., Mishra, D., & Zeeshan, S. (2021). JWES: a new pipeline for whole genome/exome sequence data processing, management, and gene-variant discovery, annotation, prediction, and genotyping. FEBS open bio, 11(9), 2441–2452. 10.1002/2211-5463.13261

12. Danecek, P., Auton, A., Abecasis, G., Albers, C. A., Banks, E., DePristo, M. A., Handsaker, R. E., Lunter, G., Marth, G. T., Sherry, S. T., McVean, G., Durbin, R., & 1000 Genomes Project Analysis Group (2011). The variant call format and VCFtools. Bioinformatics (Oxford, England), 27(15), 2156–2158. 10.1093/bioinformatics/btr330

13. Martin, F. J., Amode, M. R., Aneja, A., Austine-Orimoloye, O., Azov, A. G., Barnes, I., Becker, A., Bennett, R., Berry, A., Bhai, J., Bhurji, S. K., Bignell, A., Boddu, S., Branco Lins, P. R., Brooks, L., Ramaraju, S. B., Charkhchi, M., Cockburn, A., Da Rin Fiorretto, L., Davidson, C., … Flicek, P. (2023). Ensembl 2023. Nucleic acids research, 51(D1), D933–D941. 10.1093/nar/gkac958

14. McCarthy, D. J., Humburg, P., Kanapin, A., Rivas, M. A., Gaulton, K., Cazier, J. B., & Donnelly, P. (2014). Choice of transcripts and software has a large effect on variant annotation. Genome medicine, 6(3), 26. 10.1186/gm543

15. Sherry, S. T., Ward, M. H., Kholodov, M., Baker, J., Phan, L., Smigielski, E. M., & Sirotkin, K. (2001). dbSNP: the NCBI database of genetic variation. Nucleic acids research, 29(1), 308–311. 10.1093/nar/29.1.308

16. Liu, X., Li, C., Mou, C., Dong, Y., & Tu, Y. (2020). dbNSFP v4: a comprehensive database of transcript-specific functional predictions and annotations for human nonsynonymous and splice-site SNVs. Genome medicine, 12(1), 103. 10.1186/s13073-020-00803-9

17. Rentzsch, P., Witten, D., Cooper, G. M., Shendure, J., & Kircher, M. (2019). CADD: predicting the deleteriousness of variants throughout the human genome. Nucleic acids research, 47(D1), D886–D894. 10.1093/nar/gky1016

18. Ionita-Laza, I., McCallum, K., Xu, B., & Buxbaum, J. D. (2016). A spectral approach integrating functional genomic annotations for coding and noncoding variants. Nature genetics, 48(2), 214–220. 10.1038/ng.3477

19. Landrum, M. J., Lee, J. M., Benson, M., Brown, G. R., Chao, C., Chitipiralla, S., Gu, B., Hart, J., Hoffman, D., Jang, W., Karapetyan, K., Katz, K., Liu, C., Maddipatla, Z., Malheiro, A., McDaniel, K., Ovetsky, M., Riley, G., Zhou, G., Holmes, J. B., … Maglott, D. R. (2018). ClinVar: improving access to variant interpretations and supporting evidence. Nucleic acids research, 46(D1), D1062–D1067. 10.1093/nar/gkx1153

20. Ahmed Z. (2022). Precision medicine with multi-omics strategies, deep phenotyping, and predictive analysis. Progress in molecular biology and translational science, 190(1), 101–125. 10.1016/bs.pmbts.2022.02.002

21. Mhatre, I., Abdelhalim, H., Degroat, W., Ashok, S., Liang, B. T., & Ahmed, Z. (2023). Functional mutation, splice, distribution, and divergence analysis of impactful genes associated with heart failure and other cardiovascular diseases. Scientific reports, 13(1), 16769. 10.1038/s41598-023-44127-1

22. Venkat, V., Abdelhalim, H., DeGroat, W., Zeeshan, S., & Ahmed, Z. (2023). Investigating genes associated with heart failure, atrial fibrillation, and other cardiovascular diseases, and predicting disease using machine learning techniques for translational research and precision medicine. Genomics, 115(2), 110584. 10.1016/j.ygeno.2023.110584

23. Patel, K. K., Venkatesan, C., Abdelhalim, H., Zeeshan, S., Arima, Y., Linna-Kuosmanen, S., & Ahmed, Z. (2023). Genomic approaches to identify and investigate genes associated with atrial fibrillation and heart failure susceptibility. Human genomics, 17(1), 47. 10.1186/s40246-023-00498-0

24. Dina, C., Meyre, D., Gallina, S., Durand, E., Körner, A., Jacobson, P., Carlsson, L. M., Kiess, W., Vatin, V., Lecoeur, C., Delplanque, J., Vaillant, E., Pattou, F., Ruiz, J., Weill, J., Levy-Marchal, C., Horber, F., Potoczna, N., Hercberg, S., Le Stunff, C., … Froguel, P. (2007). Variation in FTO contributes to childhood obesity and severe adult obesity. Nature genetics, 39(6), 724–726. 10.1038/ng2048

25. Price, R. A., Li, W. D., & Zhao, H. (2008). FTO gene SNPs associated with extreme obesity in cases, controls and extremely discordant sister pairs. BMC medical genetics, 9, 4. 10.1186/1471-2350-9-4

26. Wellcome Trust Case Control Consortium (2007). Genome-wide association study of 14,000 cases of seven common diseases and 3,000 shared controls. Nature, 447(7145), 661–678. 10.1038/nature05911

27. Hinohara, K., Nakajima, T., Takahashi, M., Hohda, S., Sasaoka, T., Nakahara, K. I., Chida, K., Sawabe, M., Arimura, T., Sato, A., Lee, B. S., Ban, J. M., Yasunami, M., Park, J. E., Izumi, T., & Kimura, A. (2008). Replication of the association between a chromosome 9p21 polymorphism and coronary artery disease in Japanese and Korean populations. Journal of human genetics, 53(4), 357–359. 10.1007/s10038-008-0248-4

28. Samani, N. J., Erdmann, J., Hall, A. S., Hengstenberg, C., Mangino, M., Mayer, B., Dixon, R. J., Meitinger, T., Braund, P., Wichmann, H. E., Barrett, J. H., König, I. R., Stevens, S. E., Szymczak, S., Tregouet, D. A., Iles, M. M., Pahlke, F., Pollard, H., Lieb, W., Cambien, F., … WTCCC and the Cardiogenics Consortium (2007). Genomewide association analysis of coronary artery disease. The New England journal of medicine, 357(5), 443–453. 10.1056/NEJMoa072366

29. Silander, K., Tang, H., Myles, S., Jakkula, E., Timpson, N. J., Cavalli-Sforza, L., & Peltonen, L. (2009). Worldwide patterns of haplotype diversity at 9p21.3, a locus associated with type 2 diabetes and coronary heart disease. Genome medicine, 1(5), 51. 10.1186/gm51

30. Shen, G. Q., Rao, S., Martinelli, N., Li, L., Olivieri, O., Corrocher, R., Abdullah, K. G., Hazen, S. L., Smith, J., Barnard, J., Plow, E. F., Girelli, D., & Wang, Q. K. (2008). Association between four SNPs on chromosome 9p21 and myocardial infarction is replicated in an Italian population. Journal of human genetics, 53(2), 144–150. 10.1007/s10038-007-0230-6

31. Shen, G. Q., Li, L., Rao, S., Abdullah, K. G., Ban, J. M., Lee, B. S., Park, J. E., & Wang, Q. K. (2008). Four SNPs on chromosome 9p21 in a South Korean population implicate a genetic locus that confers high cross-race risk for development of coronary artery disease. Arteriosclerosis, thrombosis, and vascular biology, 28(2), 360–365. 10.1161/ATVBAHA.107.157248

32. Musunuru, K., Strong, A., Frank-Kamenetsky, M., Lee, N. E., Ahfeldt, T., Sachs, K. V., Li, X., Li, H., Kuperwasser, N., Ruda, V. M., Pirruccello, J. P., Muchmore, B., Prokunina-Olsson, L., Hall, J. L., Schadt, E. E., Morales, C. R., Lund-Katz, S., Phillips, M. C., Wong, J., Cantley, W., … Rader, D. J. (2010). From noncoding variant to phenotype via SORT1 at the 1p13 cholesterol locus. Nature, 466(7307), 714–719. 10.1038/nature09266

33. Cai, S., Zhang, J., Zhou, C., Shi, W., Gao, Y., Chang, R., Tan, H., Wang, Q., Ye, X., Cao, Q., Zhou, Q., Yang, P., & Hu, J. (2022). Association of CDK6 gene polymorphisms with Behcet’s disease in a Han Chinese population. Experimental eye research, 223, 109203. 10.1016/j.exer.2022.109203

34. DeGroat, W., Abdelhalim, H., Peker, E., Sheth, N., Narayanan, R., Zeeshan, S., … Ahmed, Z. (2024). Multimodal AI/ML for discovering novel biomarkers and predicting disease using multi-omics profiles of patients with cardiovascular diseases. bioRxiv, 2024.08.07.607041. 10.1101/2024.08.07.607041

35. Ahmed, Z., Wan, S., Zhang, F., & Zhong, W. (2024). Artificial intelligence for omics data analysis. BMC Methods, 1, 4. 10.1186/s44330-024-00004-5

36. DeGroat, W., Mendhe, D., Bhusari, A., Abdelhalim, H., Zeeshan, S., & Ahmed, Z. (2023). IntelliGenes: a novel machine learning pipeline for biomarker discovery and predictive analysis using multi-genomic profiles. Bioinformatics (Oxford, England), 39(12), btad755. 10.1093/bioinformatics/btad755

37. Narayanan, R., DeGroat, W., Mendhe, D., Abdelhalim, H., & Ahmed, Z. (2024). IntelliGenes: Interactive and user-friendly multimodal AI/ML application for biomarker discovery and predictive medicine. Biology methods & protocols, 9(1), bpae040. 10.1093/biomethods/bpae040

