## Supplementary Material 1 for "*VAREANT*: a bioinformatics application for gene variant reduction and annotation"

*VAREANT*: Users Guide

#### Manuscript Title

### 1. Overview

Analyzing variants in the human genome via Genome-Wide Association Studies (GWAS) holds promise in improving our understanding of disease causes and treatments [1]. A primary challenge with genomic analysis lies in its inherently large volume, requiring excessive computation. Identifying relevant information from these genomic datasets is necessary to design high-quality yet efficient data analyses. VAriant REduction and ANnoTation (*VAREANT*) is a powerful bioinformatic tool to filter, annotate, and extract relevant genomic data from variant datasets. *VAREANT* has been validated in two case-studies involving patients with heart failure (HF) and atrial fibrillation (AF). The following sections provide a detailed description of *VAREANT* and its usage.

#### 1.1 Key Features of *VAREANT*:

- **Customizable Filtering:** *VAREANT* supports a diverse set of user-driven filtering criteria to create a highly tailored dataset. Variants can be filtered by Ensembl IDs, RS numbers, or other relevant criteria.
- **Public Database Variant Annotation:** *VAREANT* leverages SnpEff [2] to annotate datasets with dbSNP [3], dbNSFP [4], and ClinVar [5] to provide comprehensive metadata about each filtered variant.
- **AI/ML Ready Preparation:** *VAREANT* efficiently extracts the variant dataset into a tabular AI/ML-ready data format, making results more immediately usable for AI/ML analysis. *VAREANT* is also capable of extracting variant datasets into a relational SQL schema to support custom data management workflows.

### 2. VAREANT

#### 2.1 Pre-processing

*VAREANT* is comprised of three standalone modules: Pre-Processing, Variant Annotation, and AI/ML Ready Data Preparation. Each module may be used independently or chained together to extract the most relevant information from a variant dataset.

The Pre-processing module is focused primarily on narrowing down a large variant dataset into a relevant subset using a series of configurable filtering criteria. For example, the user may use *VAREANT* to identify targeted variants pertaining to specific genes, as well as extract annotation-specific data from the variant file. This module is useful in reducing file sizes to minimize resources required for downstream processing. Users can identify targeted variants by genes and variant identifiers, as well as specify which annotations and sample information to retain.

##### 2.1.1 Variant Call Format

*VAREANT* supports the Variant Call Format (VCF) for representing variant datasets due to its extensibility and flexibility [6]. VCF files include metadata followed by a list of variants, each containing information about the chromosome, position, identifiers, reference and alternative alleles, quality scores, filters, annotations, and sample-specific data. *VAREANT* is compatible with the VCF specification and requires that datasets be formatted according to version 4.2.

The specification enumerates three sections present in every VCF file. The first section contains metadata about the file itself with each line beginning with '##'. This metadata might enumerate the list of filters applied to the variant file, or information about the sampling, etc. The second section is a single header line which will describe the format of the subsequent rows. This header will start with a single # and will enumerate the following columns: CHROM, POS, ID, REF, ALT, QUAL, FILTER, INFO, and optionally a FORMAT column followed by any number of sample identifiers. The first 8 columns are required and present in every dataset. Empty or missing values are indicated with a '.'.

*VAREANT* expects a VCF dataset as input for all three modules, and the Pre-processing and Annotation modules also output a VCF file.

##### 2.1.2 Configuration

The Pre-processing stage requires a configuration file (ending with a .config.ig extension) that specifies how *VAREANT* should filter the dataset. The format of this configuration file is intuitive and allows for specifying various rulesets for narrowing the input file. Each rule is enumerated on a separate line and will be in the format "Rule-Name: Value". The criteria are disjunctively applied to each variant. All rules are optional and ignored if omitted. The current version of *VAREANT* supports 8 rules that are enumerated in Table 2.1.1.

| Rule Name | Description | Example |
| --- | --- | --- |
| Retain-Info-Entries | VCF annotations present in the INFO column are structured as semicolon-separated key=value pairs. This rule allows the user to specify a subset of annotations as a pipe-separated list | Retain-Info-Entries: AF\|DP\|CSQ |

| | | |
|---|---|---|
| | of keys. Only the provided keys are retained in the filtered dataset. | |
| Retain-Variant-By-Gene-Symbol | A pipe-separated list of gene symbols. Only variants belonging to a specified gene will be retained. The rest are discarded from the final dataset. This is useful if the gene set is known a priori. | Retain-Variant-By-Gene-Symbol: BRCA2\|HBA1\|RPL21 |
| Retain-Variant-By-Ensembl-ID | A pipe-separated list of Ensembl IDs. All variants found within this list will be included. | Retain-Variant-By-Ensembl-ID: ENSG00000129562 |
| Retain-Variant-By-rsID | A pipe-separated list of RS numbers. All variants found corresponding to this list will be included in the output. | Retain-Variant-By-rsID: rs2135603633 |
| Retain-If-Passed-Filters | A VCF file may specify a list of filters that were applied to each variant, as well as enumerating which filters failed. This condition allows the user to narrow variants only if they pass all the filters specified in this pipe-separated list. The keyword "PASS" can be used to ensure only variants passing all filters are selected. | Retain-If-Passed-Filters: PASS |
| Reformat-Genotype | Every sample in a VCF file adheres to a specific format specified in the FORMAT column. This may include information like Genotype Quality, Read Depth, Genotype Phasing, etc. The VCF specification enforces that this FORMAT is a colon separated list of tokens where the first one is GT (genotype). The user may select a subset of this information to retain. | Reformat-Genotype: GT:GQ:DP |
| Minimum-Quality-Threshold | This criterion allows the user to filter for variants that meet a minimum threshold for quality. This quality score is represented in the QUAL column of the VCF file. It must be a number. | Minimum-Quality-Threshold: 500 |
| Retain-Samples | If numerous samples are present (enumerated as columns following the FORMAT column), the user may provide a set of sample IDs to retain. Any sample not included in this pipe-separated list will be removed. | Retain-Samples: SAMPLE1\|SAMPLE2\|SAMPLE3 |

Table 2.1.1: Filtering rulesets (criterion) supported by VAREANT.

#### 2.2 Variant Annotation

The second module of *VAREANT* is variant annotation. Annotation involves using public databases to retrieve relevant, variant-specific metadata. With *VAREANT*, the user is supported in annotation using three widely used annotation databases: dbSNP, dbNSFP, and ClinVar.

- **dbSNP**: dbSNP is a centralized repository of genetic variation is comprised primarily of single nucleotide polymorphisms (SNPs). It provides useful information in identifying variants.
- **dbNSFP**: dbNSFP is an aggregated database of pathogenicity scores. Pathogenicity scores provide insight into the deleteriousness of specific variants. In other words, using various metrics annotated by dbNSFP, the user can determine whether a specific variant is benign or pathogenic.
- **ClinVar:** ClinVar is an archive of variants and their significance in human disease. It provides useful data about the relationships between each variant and known diseases, which is useful as a basis for investigating gene-disease associations.

*VAREANT* uses the SnpEff utility to annotate variant datasets with any combination of these three databases. It simplifies the workflow by unifying the different annotation scripts.

#### 2.3 AI/ML Ready Data Preparation

To prepare variant data for subsequent AI/ML analysis, it is necessary to convert the dataset from a VCF format to a more AI/ML-ready data structure. *VAREANT* enables the user to extract relevant sample and variant information from the VCF file into a tabular data structure, as well as into a relational SQLite database.

- **AI/ML-Ready**: The extracted AI/ML-ready data structure is a table with variants as columns and samples as rows. This is ideal for AI/ML analysis since tabular structures are well supported by many programming libraries, and easily interpretable.
- **SQLite**: VAREANT also supports extracting relevant data into a relational data model. The data model is structured to be compatible with the JWES Entity relationship model [7]. Separate tables for variants, info (annotations), and samples are extracted. The extracted SQLite file can later be integrated into custom SQL data management workflows.

Each module may be run independently on any valid VCF file. However, we recommend filtering an input dataset using the Pre-processing module before attempting to extract into an AI/ML-Ready data format. Doing so will improve runtimes and decrease memory consumption.

### 3. Installation and Configuration

The source code necessary for installing *VAREANT* is accessible via the GitHub repository of Ahmed Lab. It is advisable to compile tool using a Python interpreter of version 2.7 or higher. The annotation phase of *VAREANT* also requires Java to be installed since SnpEff is invoked as a Java JAR executable. The user should ensure both Python 2.7+ or Java 8+ are installed on system.

#### 3.1 Preexisting modules

*VAREANT* codebase uses a few external Python modules for efficient data processing. When installing, the following modules should also be installed:

- numpy
- pandas
- python-dateutil
- pytz
- six
- tzdata

#### 3.2 Installing VAREANT

##### 3.2.1 Download from GitHub

*VAREANT* can be installed from our GitHub repository. Follow the provided steps to clone the repository:

```
# Clone VAREANT GitHub Repository

git clone https://github.com/drzeeshanahmed/Gene_VAREANT.git

# Navigate to downloaded folder

cd Gene_VAREANT/

# Verify python version >= 3.6

python --version

# Verify java version >= 8

java --version
```

##### 3.2.2 Install Python Environment

Install all Python dependencies listed in the requirements.txt file.

```
pip install -r requirements.txt
```

##### 3.2.3 Download Annotation Databases

First, download the SnpEff executable from the website (https://snpeff.blob.core.windows.net/versions/snpEff_latest_core.zip). If the user does not intend to use VAREANT's annotation feature, such as if they have their own custom annotation workflow, this step may be skipped. Once SnpEff is downloaded, locate the path of the 'SnpSift.jar' executable. This is used to annotate the VCF files using the public annotation databases.

The second step involves downloading all relevant annotation databases. VAREANT currently supports dbSNP, dbNSFP, and ClinVar. The user must download both the compressed VCF file (.vcf.gz) as well as a corresponding index file (.vcf.gz.tbi). The index file allows for more efficient processing of large VCF files. As these databases can be very large (tens of GB per database), only download the databases that are necessary for subsequent analyses. The links to download database can be found on the GitHub page.

#### 3.3 Using VAREANT

##### 3.3.1 Pre-processing

All modules are executed using the `main.py` script. This script exposes three subcommands that can be used to execute each of the three stages supported by VAREANT.

To filter a VCF dataset using VAREANT, enter the following command in your terminal window:

```
python main.py truncate \
  --input $INPUT_VCF \
  --output $OUTPUT_DIR \
  --config $CONFIG_PATH
```

- **`$INPUT_VCF`**: The path to your input dataset. For example, if the dataset is named 'dataset.vcf' and is stored in the 'datasets' folder, then the user can provide the following command line argument: `--input datasets/dataset.vcf`
- **`$OUTPUT_DIR`**: The path to the folder to save the corresponding output file to. VAREANT will generate a 'truncated.vcf' file within that folder. For example, if the user wants to save results to the 'results' folder, they may provide the option: `--output results/`. If the folder already exists, the script will first create a timestamped subfolder to avoid overwriting any files within the folder.
- **`$CONFIG_PATH`**: The path to the VAREANT configuration file that controls how VAREANT processes the input file (refer to section 2.1.2). For example, if the `.config.ig` file is stored in

the configuration folder, then the user may provide the option: `` `--config configuration/params.config.ig` ``

##### 3.3.2 Annotation

To annotate a VCF dataset using VAREANT, the user may enter the following command:

```
python main.py annotate \
  --input $INPUT_VCF \
  --output $OUTPUT_DIR \
  --snpsift $SNPSIFT_JAR \
  --dbsnp $DBSNP_PATH \
  --dbnsfp $DBNSFP_PATH \
  --clinvar $CLINVAR_PATH
```

- **`$INPUT_VCF`**: Refer to section 3.3.1
- **`$OUTPUT_DIR`**: Refer to section 3.3.1. Annotation will generate resulting output files based on which databases are provided. For example, if the user annotates their 'dataset.vcf' file with dbSNP, dbNSFP, and ClinVar, three files will be generated based on the order of annotation: dbsnp_annotated.vcf, dbnsfp_dbsnp_annotated.vcf and clinvar_dbnsfp_dbsnp_annotated.vcf.
- **`$SNPSIFT_JAR`**: The path to the SnpSift.jar executable that is used for annotation. To use the executable pre-packaged with VAREANT, the user may use the following command line argument: `` `--snpsift snpEff/SnpSift.jar` ``
- **`$DBSNP_PATH`**: This flag is optional and must be provided only if the user wants to annotate with dbSNP. The provided path must lead to the '.vcf.gz' file for dbSNP (not distributed with VAREANT). Additionally, a '.vcf.gz.tbi' file with the same name must exist on the same level. For example, if a 'dbsnp.vcf.gz' and a corresponding 'dbsnp.vcf.gz.tbi' file exists in the 'databases' folder, the user may provide the following flag: '`--dbsnp databases/dbsnp.vcf.gz`'
- **`$DBNSFP_PATH`**: This flag is also required only if the user wishes to annotate with dbNSFP (must be downloaded separately). The dbNSFP database is packaged as a `.txt.gz` database and a corresponding `.txt.gz.tbi` index. The usage is identical to $DBSNP_PATH
- **`$CLINVAR_PATH`**: This argument is required only if the user wishes to annotate with ClinVar (must be downloaded separately). The usage is identical to $DBSNP_PATH

##### 3.3.3 AI/ML-Ready Data Preparation

The following command may be used to extract a VCF file into an AI/ML ready data format.

```
python main.py extract \
  --input $INPUT_VCF \
  --output $OUTPUT_DIR
```

- **`$INPUT_VCF`**: Refer to section 3.3.1
- **`$OUTPUT_DIR`**: Refer to section 3.3.1. This extraction phase will generate two resulting datasets: a variant_db.sqlite file and a cigt_matrix.cigt.csv file. The cigt_matrix.cigt.csv file represents a tabular matrix containing the different variants per sample. This matrix is easily used by AI/ML models for predictive analysis. The variant_db.sqlite file represents a relational structure (adhering to the JWES schema). This can be integrated with existing SQL workflows or transferred to a different relational database.

For convenience, VAREANT exposes a single command that can be used to chain all three modules sequentially. This can be run using `python main.py all` and providing the sample flags as described in sections 3.3.1, 3.3.2, and 3.3.3. More details may be found in the GitHub repository.

### 4. Requirements

*VAREANT* was designed to excel in low resource environments. It is also fully compatible with all the major operating systems including MacOS, Windows, and Linux, as well as in high-performance computing systems. The Pre-processing phase excels in multi-processor environments but is still effective on single core hardware. Variant Annotation, which uses SnpEff, excels on environments with adequate memory. AI/ML-Ready Data Preparation also performs well in environments with sufficient memory capacity.

#### 4.1 Hardware Requirements

For best performance, we recommend having at least the following hardware resources.

- Minimum Processor: Single-Core CPU
- Minimum Memory: 8GB RAM
- Storage: Adequate disk space to accommodate both input, output, all the required databases for variant annotation, as well as the *VAREANT* source code itself.

### References

1. Pang T. (2002). The impact of genomics on global health. American journal of public health, 92(7), 1077–1079. https://doi.org/10.2105/ajph.92.7.1077
2. Cingolani, P., Platts, A., Wang, leL., Coon, M., Nguyen, T., Wang, L., Land, S. J., Lu, X., & Ruden, D. M. (2012). A program for annotating and predicting the effects of single nucleotide polymorphisms, SnpEff: SNPs in the genome of Drosophila melanogaster strain w1118; iso-2; iso-3. Fly, 6(2), 80–92. https://doi.org/10.4161/fly.19695
3. Sherry, S. T., Ward, M. H., Kholodov, M., Baker, J., Phan, L., Smigielski, E. M., & Sirotkin, K. (2001). dbSNP: the NCBI database of genetic variation. Nucleic acids research, 29(1), 308–311. https://doi.org/10.1093/nar/29.1.308
4. Liu, X., Li, C., Mou, C., Dong, Y., & Tu, Y. (2020). dbNSFP v4: a comprehensive database of transcript-specific functional predictions and annotations for human nonsynonymous and splice-site SNVs. Genome medicine, 12(1), 103. https://doi.org/10.1186/s13073-020-00803-9
5. Landrum, M. J., Lee, J. M., Benson, M., Brown, G. R., Chao, C., Chitipiralla, S., Gu, B., Hart, J., Hoffman, D., Jang, W., Karapetyan, K., Katz, K., Liu, C., Maddipatla, Z., Malheiro, A., McDaniel, K., Ovetsky, M., Riley, G., Zhou, G., Holmes, J. B., … Maglott, D. R. (2018). ClinVar: improving access to variant interpretations and supporting evidence. Nucleic acids research, 46(D1), D1062–D1067. https://doi.org/10.1093/nar/gkx1153
6. Danecek, P., Auton, A., Abecasis, G., Albers, C. A., Banks, E., DePristo, M. A., Handsaker, R. E., Lunter, G., Marth, G. T., Sherry, S. T., McVean, G., Durbin, R., & 1000 Genomes Project Analysis Group (2011). The variant call format and VCFtools. Bioinformatics (Oxford, England), 27(15), 2156–2158. https://doi.org/10.1093/bioinformatics/btr330
7. Ahmed, Z., Renart, E. G., Mishra, D., & Zeeshan, S. (2021). JWES: a new pipeline for whole genome/exome sequence data processing, management, and gene-variant discovery, annotation, prediction, and genotyping. FEBS open bio, 11(9), 2441–2452. https://doi.org/10.1002/2211-5463.13261

### Acknowledgments

We appreciate great support by the Department of Medicine, Robert Wood John-son Medical School; Rutgers Institute for Health, Health Care Policy, and Aging Research; and Rutgers Health, at Rutgers, The State University of New Jersey.
