## Supplementary Material 2 for "*VAREANT*: a bioinformatics application for gene variant reduction and annotation"

*VAREANT*: Patient Demographic Data and Performance Metrics

#### Manuscript Title

### Patient Demographics

The case studies outlined in the manuscript were performed on a curated cohort of 96 samples, aggregated from 2 separate cardiovascular disease (CVD) studies. The following two tables (Table 1A, Table 1B) outline the demographic information (e.g. gender, age) for each patient.

| # | ID | Gender | Age |
|---|---|---|---|
| 1 | 1065 | Female | 51 |
| 2 | 1071 | Female | 52 |
| 3 | 1064 | Female | 54 |
| 4 | 1114 | Female | 54 |
| 5 | 1097 | Female | 57 |
| 6 | 1108 | Female | 57 |
| 7 | 1075 | Female | 59 |
| 8 | 1082 | Female | 59 |
| 9 | 1116 | Female | 63 |
| 10 | 1069 | Female | 65 |
| 11 | 1088 | Female | 65 |
| 12 | 1084 | Female | 69 |
| 13 | 1087 | Female | 69 |
| 14 | 1093 | Female | 70 |
| 15 | 1105 | Female | 71 |
| 16 | 1058 | Female | 72 |
| 17 | 1078 | Female | 72 |
| 18 | 1074 | Female | 81 |
| 19 | 1111 | Female | 86 |
| 20 | 1073 | Female | 89 |
| 21 | 1072 | Female | 91 |

| | | | |
|---|---|---|---|
| 22 | 1076 | Male | 45 |
| 23 | 1089 | Male | 55 |
| 24 | 1070 | Male | 57 |
| 25 | 1081 | Male | 57 |
| 26 | 1060 | Male | 58 |
| 27 | 1096 | Male | 59 |
| 28 | 1113 | Male | 60 |
| 29 | 1067 | Male | 62 |
| 30 | 1092 | Male | 62 |
| 31 | 1085 | Male | 64 |
| 32 | 1094 | Male | 64 |
| 33 | 1101 | Male | 64 |
| 34 | 1086 | Male | 65 |
| 35 | 1063 | Male | 66 |
| 36 | 1095 | Male | 66 |
| 37 | 1117 | Male | 66 |
| 38 | 1062 | Male | 67 |
| 39 | 1099 | Male | 67 |
| 40 | 1115 | Male | 67 |
| 41 | 1061 | Male | 70 |
| 42 | 1068 | Male | 70 |
| 43 | 1090 | Male | 70 |
| 44 | 1102 | Male | 71 |
| 45 | 1112 | Male | 72 |
| 46 | 1077 | Male | 73 |

| | | | |
|---|---|---|---|
| 47 | 1104 | Male | 73 |
| 48 | 1109 | Male | 75 |
| 49 | 1091 | Male | 77 |
| 50 | 1059 | Male | 79 |
| 51 | 1106 | Male | 79 |
| 52 | 1103 | Male | 80 |
| 53 | 1110 | Male | 80 |
| 54 | 1100 | Male | 81 |
| 55 | 1066 | Male | 82 |
| 56 | 1098 | Male | 83 |
| 57 | 1107 | Male | 84 |
| 58 | 1083 | Male | 85 |
| 59 | 1080 | Male | 86 |
| 60 | 1118 | Male | 88 |
| 61 | 1079 | Male | 92 |

**Table 1A. Patient Demographics (Cohort A)**. This table enumerates sample ID, gender, and age of 61 patients [1], sorted by gender then by age. There are 21 females (colored orange), and 40 males (colored blue).

| # | ID | Gender | Age |
|---|---|---|---|
| 1 | BR2-1228 | Female | 56 |
| 2 | BR2-1154 | Female | 58 |
| 3 | BR2-1421 | Female | 58 |
| 4 | BR2-1094 | Female | 61 |
| 5 | BR2-1267 | Female | 69 |
| 6 | BR2-922 | Female | 72 |
| 7 | BR2-738 | Female | 73 |
| 8 | BR2-765 | Female | 74 |
| 9 | BR2-1572 | Female | 76 |
| 10 | BR2-821 | Female | 80 |
| 11 | BR2-747 | Female | 83 |
| 12 | BR2-860 | Female | 84 |
| 13 | BR2-986 | Female | 89 |
| 14 | BR2-731 | Female | 90 |
| 15 | BR2-1343 | Male | 29 |
| 16 | BR2-1506 | Male | 51 |
| 17 | BR2-975 | Male | 58 |
| 18 | BR2-1007 | Male | 58 |
| 19 | BR2-781 | Male | 62 |
| 20 | BR2-810 | Male | 62 |
| 21 | BR2-1294 | Male | 62 |
| 22 | BR2-833 | Male | 67 |
| 23 | BR2-1218 | Male | 68 |
| 24 | BR2-1366 | Male | 69 |

| | | | |
|---|---|---|---|
| 25 | BR2-1282 | Male | 70 |
| 26 | BR2-1563 | Male | 71 |
| 27 | BR2-890 | Male | 72 |
| 28 | BR2-995 | Male | 72 |
| 29 | BR2-1381 | Male | 76 |
| 30 | BR2-774 | Male | 77 |
| 31 | BR2-745 | Male | 78 |
| 32 | BR2-1273 | Male | 86 |
| 33 | BR2-1090 | Male | 94 |
| 34 | BR2-1095 | Male | 96 |
| 35 | BR2-1606 | Male | 63 |

**Table 1B. Patient Demographics (Cohort B)**. This table enumerates sample ID, gender, and age of 34 patients [2], sorted by gender then by age. There are 14 females (colored orange), and 20 males (colored blue).

### *VAREANT* Performance Metrics

The following three tables (Table 2A, Table 2B, Table 2C) detail numerous statistics about the performance of each of the three modules of *VAREANT* on our custom datasets.

| | | |
|---|---|---|
| File Size (Before) | 527 MB | 2.6 GB |
| File Size (After) | 15 MB | 1.2 MB |
| # of Variants (Before) | 99777 | 4051911 |
| # of Variants (After) | 2931 | 798 |
| Duration (4 CPUs + 8 GB, Single-threaded) | 6.9 s | 96 s |
| Duration (4 CPUs + 8 GB, Multi-threaded) | 4.3 s | 28.1 s |
| Duration (12 CPUs + 32 GB, Single-threaded) | 6.8 s | 95 s |
| Duration (12 CPUs + 32 GB, Multi-threaded) | 3.9 s | 14.1 s |

**Table 2A. Performance of *VAREANT* Pre-Processing.** This table outlines performance metrics of *VAREANT's* Pre-Processing module on two variable sized datasets (527 MB, 2.6 GB). It details statistics about the dataset before filtering, as compared to after filtering. It also lists the average processing duration in 4 different hardware environments (results subject to hardware and dataset itself).

| | | |
|---|---|---|
| File Size | 527 MB | 2.6 GB |
| Duration (Without filtering) | 14 m 45 s | 3 h 37 m 11 s |
| Duration (With filtering) | 44 s | 16 s |
| File Size (Without filtering) | 579 MB | 3.7 GB |
| File Size (With filtering) | 16 MB | 1.4 MB |

**Table 2B. Performance of *VAREANT* Annotation.** This table outlines performance metrics of *VAREANT's* Annotation module on two variable sized datasets (527 MB, 2.6 GB). It details statistics about the size of the datasets and processing duration on the original unfiltered dataset, as compared to after filtering with *VAREANT*.

| | | |
|---|---|---|
| File Size | 527 MB | 2.6 GB |
| Duration (Without filtering) | 1.2 s | 0.74 s |
| Duration (With filtering) | 0.9 s | 0.7 s |
| Tabular File Size (Without filtering) | 20 MB | 11 KB |
| Tabular File Size (With filtering) | 569 KB | 3.2 KB |
| Relational DB Size (Without filtering) | 2.3 GB | 11.2 MB |
| Relational DB Size (With filtering) | 36 MB | 3.8 MB |

**Table 2C. Performance of *VAREANT* AI/ML Data Preparation.** This table outlines performance metrics of *VAREANT's* AI/ML Data Preparation module on two variable sized datasets (527 MB, 2.6 GB). It enumerates the sizes of the extracted AI/ML ready datasets, as well as the general processing duration.

### References

1. Venkat, V., Abdelhalim, H., DeGroat, W., Zeeshan, S., & Ahmed, Z. (2023). Investigating genes associated with heart failure, atrial fibrillation, and other cardiovascular diseases, and predicting disease using machine learning techniques for translational research and precision medicine. Genomics, 115(2), 110584. https://doi.org/10.1016/j.ygeno.2023.110584
2. Mhatre, I., Abdelhalim, H., Degroat, W., Ashok, S., Liang, B. T., & Ahmed, Z. (2023). Functional mutation, splice, distribution, and divergence analysis of impactful genes associated with heart failure and other cardiovascular diseases. Scientific reports, 13(1), 16769. https://doi.org/10.1038/s41598-023-44127-1

### Acknowledgments

We appreciate great support by the Department of Medicine, Robert Wood John-son Medical School; Rutgers Institute for Health, Health Care Policy, and Aging Research; and Rutgers Health, at Rutgers, The State University of New Jersey.
